# Multiple effects of TNFα inhibitors on the development of the adaptive immune response after SARS-CoV-2 vaccination

**DOI:** 10.1101/2022.07.01.22277143

**Authors:** Ulf Martin Geisen, Ruben Rose, Franziska Neumann, Maria Ciripoi, Lena Vullriede, Hayley M Reid, Dennis Kristopher Berner, Federico Bertoglio, Paula Hoff, Michael Hust, Ann Carolin Longardt, Thomas Lorentz, Gabriela Rios Martini, Carina Saggau, Jan Henrik Schirmer, Maren Schubert, Melike Sümbül, Florian Tran, Mathias Voß, Rainald Zeuner, Peter J Morrison, Petra Bacher, Helmut Fickenscher, Sascha Gerdes, Matthias Peipp, Stefan Schreiber, Andi Krumbholz, Bimba Franziska Hoyer

**Affiliations:** Medical Department I, Rheumatology and Clinical Immunology, University Medical Center Schleswig-Holstein Campus Kiel, Kiel, Germany; Institute for Infection Medicine, Christian-Albrecht University of Kiel and University Medical Center Schleswig-Holstein, Campus Kiel, Kiel, Germany; Labor Dr. Krause und Kollegen MVZ GmbH, Kiel, Germany; Institute of Biochemistry, Biotechnology, and Bioinformatics, Department of Biotechnology, Technische Universität Braunschweig, Braunschweig, Germany; Department of Rheumatology, Endokrinologikum-Gruppe, Berlin, Germany; Department of Pediatrics, University Medical Center Schleswig-Holstein Campus Kiel, Kiel, Germany; Institute of Immunology, University Medical Center Schleswig-Holstein Campus Kiel, Kiel, Germany; Institute of Clinical Molecular Biology, Christian-Albrecht University of Kiel, Kiel, Germany; Department for Dermatology, University Medical Center Schleswig-Holstein Campus Kiel, Kiel, Germany; Department for Internal Medicine I, University Medical Center Schleswig-Holstein Campus Kiel; Division of Antibody-Based Immunotherapy, Department of Internal Medicine II, University Medical Center Schleswig-Holstein Campus Kiel, Kiel, Germany

**Keywords:** COVID19, Tumor Necrosis Factor Inhibitors, Vaccination, Autoimmune Diseases

## Abstract

**Objectives:** The humoral immune response to SARS-CoV-2 vaccination in patients with chronic inflammatory disease (CID) declines more rapidly with TNFα inhibition. Furthermore, the efficacy of current vaccines against Omicron variants of concern (VOC) including BA.2 is limited. Alterations within immune cell populations, changes in IgG affinity and the ability to neutralise a pre-VOC strain and the BA.2 virus were investigated in these at-risk patients.

**Methods:** Serum levels of anti-SARS-CoV-2 IgG, IgG avidity and neutralising antibodies (NA) were determined in anti-TNFα patients (n=10) and controls (n=24 healthy individuals; n=12 patients under other disease-modifying anti-rheumatic drugs, oDMARD) before and after the second and third vaccination by ELISA, immunoblot and live virus neutralisation assay. SARS-CoV-2-specific B-and T cell subsets were analysed by multicolour flow cytometry.

**Results:** IgG avidity and anti-pre-VOC NA titres decreased faster in anti-TNFα recipients than in controls 6 months after the second vaccination (healthy individuals: avidity: p≤0.0001; NA: p=0.0347; oDMARDs: avidity: p=0.0012; NA: p=0.0293). Total plasma cell counts were increased in anti-TNFα patients (Healthy individuals: p=0.0344; oDMARDs: p=0.0254), whereas absolute numbers of SARS-CoV-2-specific cells were comparable 7 days after vaccination. These patients had lower BA.2 NA titres compared to both other groups, even after the third vaccination.

**Conclusions:** We show a reduced SARS-CoV-2 neutralising capacity in patients under TNFα blockade. In this cohort, the plasma cell response appears to be less specific and show stronger bystander activation. While these effects were observable after the first two vaccinations and with older VOC, the differences in responses to BA.2 were magnified.

**What is already known on this topic:** Patients with chronic inflammatory diseases treated with TNFα inhibitors show a greater decrease in SARS-CoV-2 IgG 6 months after the second vaccination than patients taking oDMARDs and healthy individuals.

**What this study adds:** Antibodies from patients taking TNFα blockers have a lower SARS-CoV-2 neutralising capacity and maturity. Plasma cells from these patients exhibit less specific immune reaction. SARS-CoV-2-specific T cells are less activated. Neutralisation against BA.2 is drastically reduced even after the third vaccination.

**How this study might affect research, practice or policy:** This study emphasizes the need to protect vulnerable groups such as patients using TNF inhibitors. They could benefit from Omicron-adapted vaccination, but most likely they need to be protected by additional means other than vaccination.

## Introduction

The current SARS-CoV-2 pandemic poses a particular challenge for patients with chronic inflammatory disease (CID) receiving immunosuppressive therapies. For example, certain immunosuppressive therapies/pharmaceuticals (e.g., B cell depleting therapies, antimetabolites such as methotrexate, high-dose corticosteroids) are known to interfere with SARS-CoV-2 vaccine efficacy.(1) However, long-term data from this population on immune response to the vaccines are lacking.

Previously, we found that CID patients under tumour necrosis factor alpha (TNFα) inhibiting therapy initially showed a largely normal, albeit slightly delayed, immune response to SARS-CoV-2 mRNA vaccines which was followed by a rapid decline of anti-spike (S) and virus-neutralising antibody (NA) levels compared to patients receiving other disease modifying anti-rheumatic drugs (oDMARDs) and healthy controls.(2) While the difference in anti-S antibody levels was marginal at day 7 and absent at day 14 after the second vaccination, these patients had significantly lower anti-S IgG levels six months after vaccination. Moreover, the neutralising capacity of serum in CID patients treated with TNFα inhibitors was dramatically reduced at the sixth month after vaccination, as shown by a surrogate neutralisation assay.(3) This impairment of adaptive immunity during anti-TNFα treatment has also been confirmed by other research groups, including live virus neutralisation data using the Delta variant of concern (VOC) as antigen.(4, 5) Compared with healthy controls, anti-S IgA levels were decreased in CID patients at all time points after vaccination, suggesting impaired mucosal immunity.(3) It remains unclear what biological mechanisms lead to this impaired antibody response and whether these differences indicate generally lower immunity after vaccination compared with controls. The relationship between B cells and T cells during SARS-CoV-2 vaccination is not fully understood, as humoral and T cell immunity appear to depend on B cell counts before vaccination.(6) In addition, data from immunocompromised kidney transplant patients show that T cell activity after vaccination correlates with the magnitude of the antibody response,(7) while high T cell activity has been observed in B cell depleted patients after immunisation.(8)

Sera from vaccinated healthy individuals show only limited neutralisation capacity against Omicron (B.1.1.529) VOC.(9, 10) This variant, consisting of several sublineages, including BA.1 and BA.2, is considered a separate serotype that is antigenically distinct from the original Wuhan strain (designated here as wild-type, wt, or pre-VOC) and other VOCs.(11) The marked immune escape of BA.1 and BA.2 and the importance of booster vaccination for the development of NA against both sublineages have recently been demonstrated,(10) especially the need of mRNA boost immunisations for persons vaccinated with inactivated viruses.(12)

Only limited data are available on the persistence of NA against various SARS-CoV-2 lineages (including Omicron) in CID patients receiving anti-TNFα therapy after double vaccination. Virtually no data are available on the development of binding strength (avidity) of vaccine-induced IgG antibodies, which is considered an expression of their maturity and optimal epitope binding (13) (14), for this group of patients, nor are there any data on the development of cellular immunity.

The aim of this study is to clarify the influence of immunosuppressive therapy on the development of adaptive immunity after SARS-CoV-2 vaccination. To this end, the quality and quantity of SARS-CoV-2-specific B cells, plasmablasts, T cells, and antibodies were measured at different time points after the second vaccination. We report differential development of anti-BA.2 NAs after a third dose of vaccine.

## Methods

### Patient recruitment and biosampling

The study was approved by the ethics committee of the Christian-Albrecht University Kiel (D409/21). There was no patient and public involvement in conducting the study. Recruitment of patients and repetitive biosampling was performed as previously described.(2) The SAVE-CID cohort consists of 47 healthcare workers and other risk groups who received their first SARS-CoV-2 vaccination in January 2021 followed by a second vaccination 5 or 3 weeks later. Samples taken 7 days after the third vaccination were also examined in 12 patients. All patients received BNT162b2 (Comirnaty, Pfizer/BioNTech) or mRNA-1273 (Spikevax, Moderna). Biosampling and data acquisition as well as data on antibody concentration, surrogate neutralisation data and clinical characterisation of this cohort has already been published.(2, 3) The patient groups were age and gender matched resulting in mean ages of 43 (TNFα inhibitor, median: 42.5), 41.25 (Healthy Control, median: 39) and 41.63 (oDMARDs, median: 46).

### Production of SARS-CoV-2 S1 proteins

S1 domain of the S protein(GenBank: MN908947) with different tags were baculovirus-free produced in High Five cells (Thermo Fisher Scientific) by transient transfection as previously described.(15, 16) Protein purification was performed depending 1 or 5 mL column on Äkta go (Cytiva), Äkta Pure (Cytiva), or Profina System (BIO-RAD). HiTrap Fibro PrismA (Cytiva) was used as resins for Protein A purification (Fc-tagged proteins). For His-tag purification of insect cell supernatant HisTrapexcel column (Cytiva) was used. All purifications were performed according to the manufacturer’s manual. S1-HIS was further purified by SEC by a 16/600 Superdex 200 kDa pg (Cytiva).

### Isolation of peripheral blood mononuclear cells (PBMC) and Flow Cytometry

PBMCs from EDTA blood were isolated within three hours of blood collection by density gradient centrifugation (Biocoll, Bio&SELL GmbH, Feucht, Germany). Afterwards, 4×10^6^ PBMCs were incubated with his-tagged S1 protein (own protein or Euroimmun, Lübeck, Germany).(9) PBMCs were then stained with pre-mixed antibodies (CD19-PerCP-Vio-700 (REA657, Miltenyi Biotec, Bergisch-Gladbach, Germany), CD20-PE-Cio770 (REA780, Miltenyi Biotec), CD3-Pacific Blue (OKT3, Biolegend, San Diego, USA), CD14 Pacific Blue (M5E2, Biolegend), CD27-APC (M-T271, Biolegend), anti-HIS-PE (JO95-G46, Biolegend), Biolegend), HLA-DR-VioGreen (REA805, Miltenyi Biotec), CD138-BV605 MI15, Biolegend)) and analysed using a MacsQuant 16 Cytometer (Miltenyi Biotec, Bergisch-Gladbach, Germany). Secondary staining using AF-700 coupled S1-fc protein were used as gating and staining control to exclude false positive events (see figure S1 and S2 for more information). For the calculation of immune cells per blood volume, 50µL of whole blood was stained, (CD3-Pacific Blue (Biolegend), CD14-FITC (REA599, Miltenyi Biotec), CD4-PE (Vit4, Miltenyi Biotec), CD19-PerCP-Vio700 and CD45-APC-Vio770 (H130, Biolegend)), lysed, (Red blood lysis, BD) and measured on a MACSQuant 16 (Miltenyi Biotec). The cell counts per 50µL blood for each sample were used to calculate all other cell counts from the PBMC staining. Staining and measurement were performed in PBS containing 1% BSA, 0.5% EDTA and 0.1% sodium azide.

### Measurement of Antibody Secreting Cells by Three-Colour Fluorospot

PBMCs were isolated as described above and different T cell numbers were incubated for three hours in a 96-well 0.45µM PVDF Immobilon-FL membrane plate (Merck Millipore, Burlington, MS, USA) which was pre-coated with SARS-CoV-2 protein or FAB2-fragments against Immunoglobulins A, M (both Southern Biotech, Birmingham, USA), G (Jackson Immunoresearch, Cambridge, UK) and blocked with 15%. FCS in PBS. After 3 washing steps with deionised water, the wells were stained using IgA-FITC, IgG-FC-AF555 and IgM-AF647 (all Southern Biotech). Measurement was performed on a Bioreader 6000Fb equipped with Eazyreader software (Bio-SYS, Karben, Germany)

### Antigen-reactive T cell enrichment (ARTE)

The ARTE was performed as previously described (17-20). In brief, 0.5-1×10^7^ PBMCs were stimulated for 7 hours with 0.5 µg/peptide/ml SARS-CoV-2 S peptide pool (JPT, Berlin, Germany) in presence of 1 µg/ml CD40 and 1 µg/ml CD28 pure antibody (both Miltenyi Biotec, Bergisch Gladbach, Germany). 1 µg/ml Brefeldin A (Sigma Aldrich) was added for the last two hours. Cells were magnetically isolated using the CD154 MicroBead Kit (Miltenyi Biotec). After surface staining with CD4-APC-Vio770 (M-T466), CD8-VioGreen (REA734), CD14-VioGreen (REA599), CD20-VioGreen (LT20) (all Miltenyi Biotec), CD45RA-PE-Cy5 (HI100), PD-1 Brilliant Violet 605 (EH12.2H7), CCR7-Brilliant-Violet-785 (G043H7) (all BioLegend), cells were fixed, permeabilized and stained intracellular with CD154-FITC (REA238), IL-21-PE (REA1039) (both Miltenyi Biotec), IFN-γ-PerCP-Cy5.5 (4S.B3), TNFα-Brilliant-Violet-650 (MAb11), IL-10-PE-Dazzle (JES3-9D7) (all BioLegend), IL-2-BV711 (5344.111), Ki-67-Alexa Fluor 700 (B56) (both BD Biosciences). Viobility 405/520 Fixable Dye (Miltenyi Biotec) was used to exclude dead cells. Data were acquired on a LSR Fortessa (BD Bioscience, San Jose, CA, USA). CD154^+^ background cells enriched from the non-stimulated control were subtracted and frequencies of antigen-specific T cells were determined based on CD154^+^ T cells after enrichment, normalised to the total number of CD4^+^ T cells applied on the column.

### Binding strength (avidity) of SARS-CoV-2 IgG antibodies

The IgG avidity was assessed with the *recom*Line SARS-CoV-2 IgG assay on a Dynablot Plus system together with a BLOTrix reader and the recomScan software (all from Mikrogen GmbH, Neuried, Germany) as reported previously.(21) This immunoblot consists of a nitrocellulose strip separately carrying recombinant nucleocapsid protein (NP) and the S1-and RBD-subunits of the S protein. Binding of IgG to these SARS-CoV-2 antigens in presence or absence of avidity reagent was automatically measured and assigned to four categories: no avidity detectable (=0), low avidity (=1), intermediate avidity (=2) and high (=3) avidity.(22)

### Measurement of neutralising antibodies against a pre-VOC strain and a BA.2 strain

Sera were tested in triplicate using a Vero cell-based live virus neutralisation test (cVNT) in 96-well format under biosafety level 3 conditions, as previously reported.(10, 22) In brief, sera were diluted 1:10 to 1:1280 in cell culture medium free of fetal calf serum. As antigens for the cVNT, we used either 50 plaque-forming units per well of a B.1 strain (pre-VOC of 2020) or an Omicron BA.2 strain, which we had previously isolated (23) and characterised by whole-genome sequencing.(10, 23) After four (pre-VOC) or six (BA.2) days of incubation, cells were fixed by addition of paraformaldehyde and stained with an aqueous crystal violet methanol solution. Serum dilutions (titres) > 1:10 that prevented the formation of a cytopathic effect in ≥2 wells were considered to contain neutralising antibodies (NA); if no exact titre could be given, the geometric mean of the two adjacent titres was calculated.(10, 22)

### Data analysis and statistics

Flow cytometry data was analysed using FlowJo v10 (BD Bioscience). Statistical analyses and graphs were prepared using RStudio (version 2022.02.0+443) and Prism 8 (GraphPad Software, LLC). For analysis of differences between the groups, Kruskal-Wallis Test and non-parametric pairwise comparisons were performed.

## Results

### Anti-S-IgG-antibody concentrations, IgG avidity and neutralisation efficacy decrease 6 months after vaccination

Under TNFα inhibitor therapy, anti-S-IgG levels after the second immunisation were significantly lower than in patients receiving oDMARDs or in healthy controls (figure 1a). None of the subjects showed anti-NP IgG reactivity, making infection breakthrough unlikely (data not shown). The IgG avidity and neutralisation capacity against the pre-VOC strain were high in all tested groups 14 days after second vaccination (avidity: median avidity index (MAI) = 3; NA: geometric mean titres (GMT)=1:98–1:234)(figure 1B, C). Six months after second vaccination, IgG avidity and pre-VOC NA titres significantly decreased in TNFα-inhibitor treated vaccinees (n=8; avidity: MAI=1.25; NA: GMT=1:2) compared to patients receiving oDMARDs (n=7; avidity: MAI=3, p=0.0012; NA: GMT=1:38, p=0.0293) and healthy controls (n=12; avidity: MAI=3, p ≤0.0001; NA: GMT=1:25, p=0.0347) (figure 1A). Relative to the pre-VOC strain, anti-BA.2 NA titres were significantly lower than against the pre-VOC strain in all three groups at day 14 after the second vaccination (GMT=1:2-1:4 vs. GMT=1:98-1:234; p=0.0001-0.0072) and were not detected after six months (GMT<1:10, figure 1D). At 7 days after the third vaccination, anti-BA.2 NAs were detectable in all subjects except patients taking a TNFα blocker (n=4 per group; GMT=1:62-1:95 vs. GMT=1:3). At this time point, IgG avidity was high in all subjects except one patient receiving anti-TNFα treatment.

**Figure 1:**
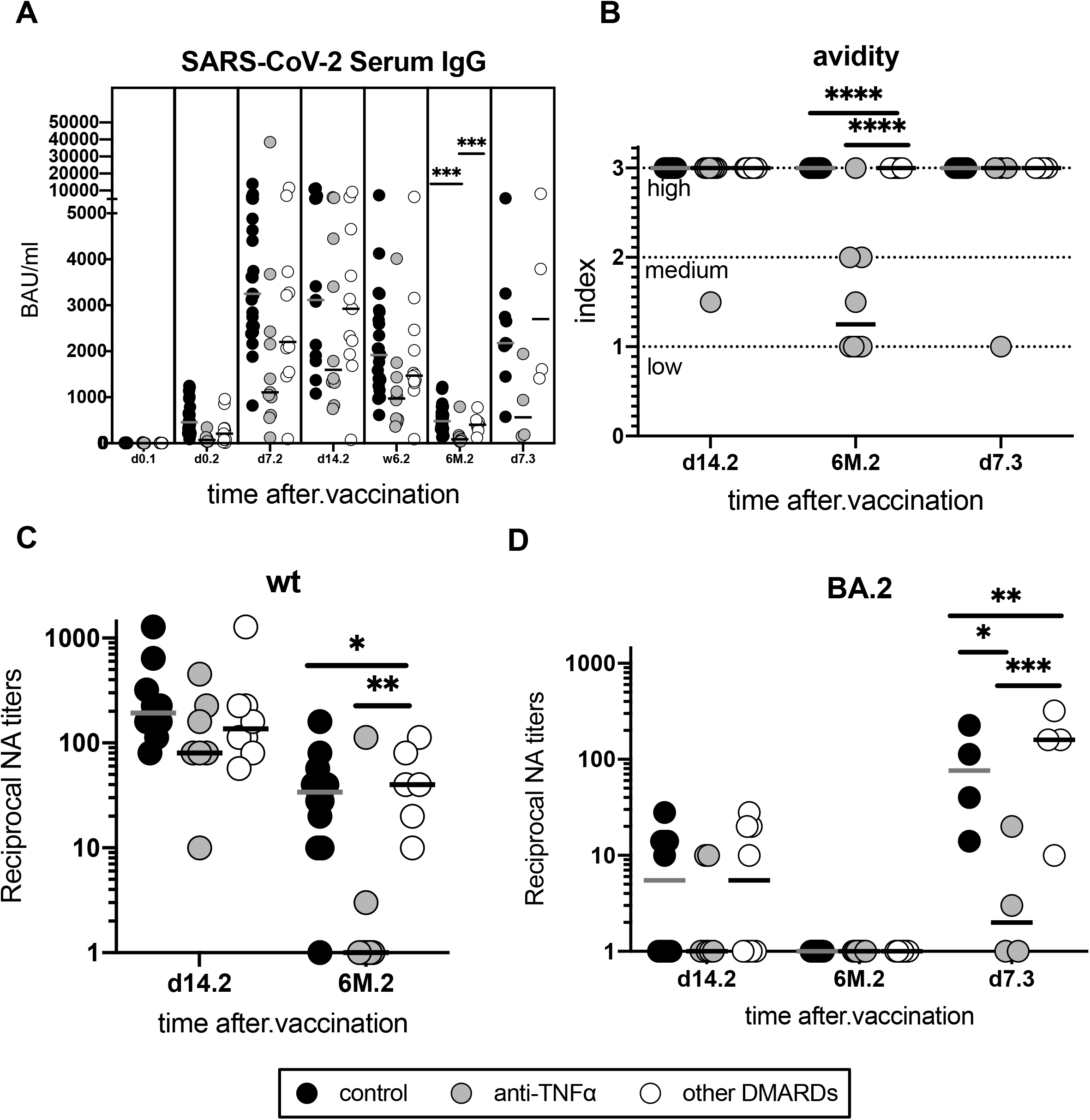
**A** Patients using TNFα blockers show reduced antibody serum levels, avidiy and neutralisation at different timepoints. Serum IgG antibody against SARS-CoV-2 Spike 1 subunit at different timepoints as single values per patient. Lines indicate the median. **B** IgG avidity indices of anti-S IgGs of SARS-CoV-2. **C** Neutralising antibodies against the wild-type (wt) and **D** BA.2 variant of SARS-CoV-2 at 14 days and six months after the second vaccination. For BA.2 and IgG avidity, a time point 7 days after the third vaccination was added (n=4 per group); values are given as individual values and median. Statistical differences: Kruskal-Wallis test with Dunn’s post hoc test, significant differences are indicated as *\*P<0*.*05, **P<0*.*01, ***P<0*.*001, ****P<0*.*0001*

### Plasma cell populations are altered in patients using TNFα blockers

Patients under TNFα inhibiting therapy showed higher numbers of plasmablasts in the peripheral blood 7 days after the second vaccination (median: 9.153cells/µL) compared to patients receiving oDMARDs (median: 2.205cells/µL, p=0.0254) (figure 2A) and healthy controls (2.657 cells/µL, p=0.0344).

**Figure 2:**
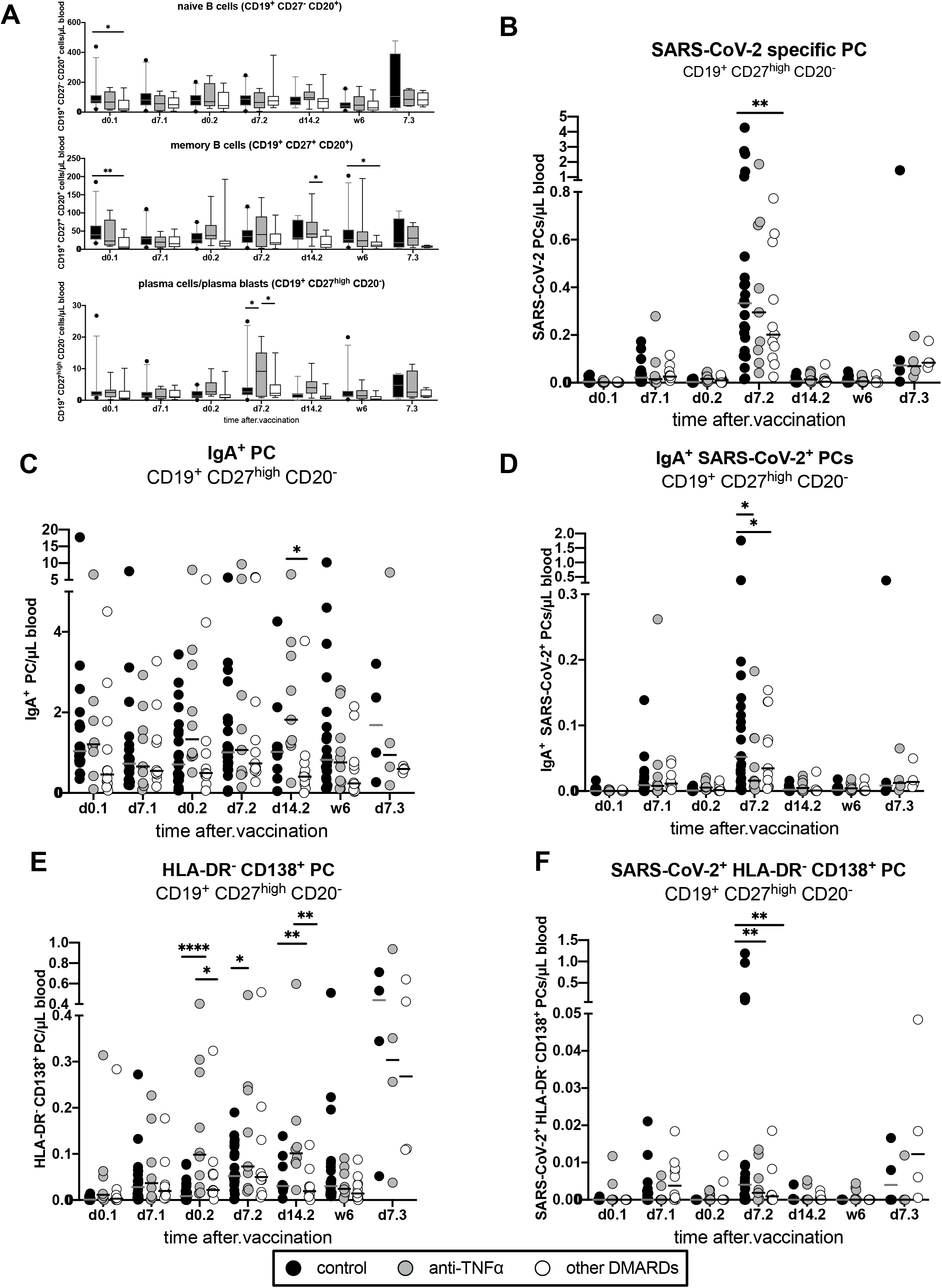
Patients using TNFα blockers show higher numbers of overall plasmablasts and SARS-CoV-2 specific plasmablasts 7 days after the second vaccination. **A** Flow cytometry analysis of B cell subsets at different timepoints after a vaccination against SARS-CoV-2. Values are shown as cells per microlitre of blood. Boxes and whiskers indicate median and 95%CI. **B** SARS-CoV-2 specific plasmablasts. IgA^+^ plasmablasts **C** and SARS-CoV-2 specific **D** IgA^+^ positive plasmablasts. **E** and **F**: CD138^+^ unspecific and SARS-CoV-2 specific plasmablasts which resemble mature plasma cells. If not indicated otherwise, cell counts per microlitre blood are shown as single values with median. Statistical differences: Kruskal-Wallis test with Dunn’s post hoc test, significant differences are indicated as *P<0.05, \*\**P*<0.01, \*\*\*\**P*<0.0001

When comparing SARS-CoV-2 specific plasma cells at the same timepoint, no differences between patients on TNFα blockade (median 0.295 cells/µL) and healthy controls (median 0.333 cells/µL) were detected, while patients treated with oDMARDs other than TNFα inhibitors had significantly lower cell numbers compared to healthy controls (median: 0.204 cells/µL; p=0.0015) (figure 2B).

Anti-TNFα treated patients generally displayed more peripheral blood IgA plasma cells than controls and patients receiving oDMARDs (figure 2C). This trend became significant on day 14 after vaccination for patients under oDMARDs (median 0.404 cells/µL) and TNFα blockers (median 1.818 cells/µL; p=0.0397).

With regards to SARS-CoV-2-specific IgA-plasma cells, counts were lower in the anti-TNFα group on day 7 after the second vaccination (median: 0.016 cells/µL) compared to healthy controls (median: 0.052 cells/µL; p=0.0203) and patients treated with oDMARDs (median:0.035 cells/µL; p=0.0299) (figure 1D).

Overall numbers of mature circulating CD138^+^ plasma cells were higher in patients on TNFα blockade (median: 0.101 cells/µL) than in healthy controls (median: 0.031 cells/µL; p=0.0055) and patients treated with oDMARDs (median: 0.019 cells/µL; p=0.0015) on day 14 after second vaccination (figure 2E). Differences at time points between the first and the second vaccination were not significant. We were not able to detect differences in the number of SARS-CoV-2 specific plasma cells. However, on day 7 after second vaccination, healthy controls had higher numbers of circulating CD138^+^ plasma cells in the peripheral blood (median: 0.004 cells/µL) than patients using TNFα inhibitors (median: 0.002 cells/µL; p=0.0055) or receiving oDMARDs (median: 0.001 cells/µL; p=0.0026) (figure 2F).

### Anti-TNFα treatment does not change the number of SARS-CoV-2 specific antibody secreting cells

Relative to the other two sample groups, anti-TNFα patients showed a pronounced increase in SARS-CoV-2 specific antibody secreting cells (ASCs) of the IgM isotype at day 7 post second vaccination, while total numbers of ASC of all other isotypes remained comparable between groups (figure 3A). Analysing spot size in our Fluorospot assays as a surrogate for the amount of secreted antibody per cell, both patient groups generally displayed larger spot sizes suggesting increased antibody secretion per ASC (figure 3B). These differences were not significant except for IgA (p=0.0026 for TNF vs. Healthy Control and p=0.001 for oDMARDs vs. Healthy Control). No differences were detected between the two patient groups. The number of ASCs correlated well with SARS-CoV-2 serum IgG levels and the number of SARS-CoV-2 positive plasmablasts at the same timepoint (figure 3C-D). No antigen specific plasma cells were detected in the blood of any participant before the first vaccination (data not shown).

**Figure 3:**
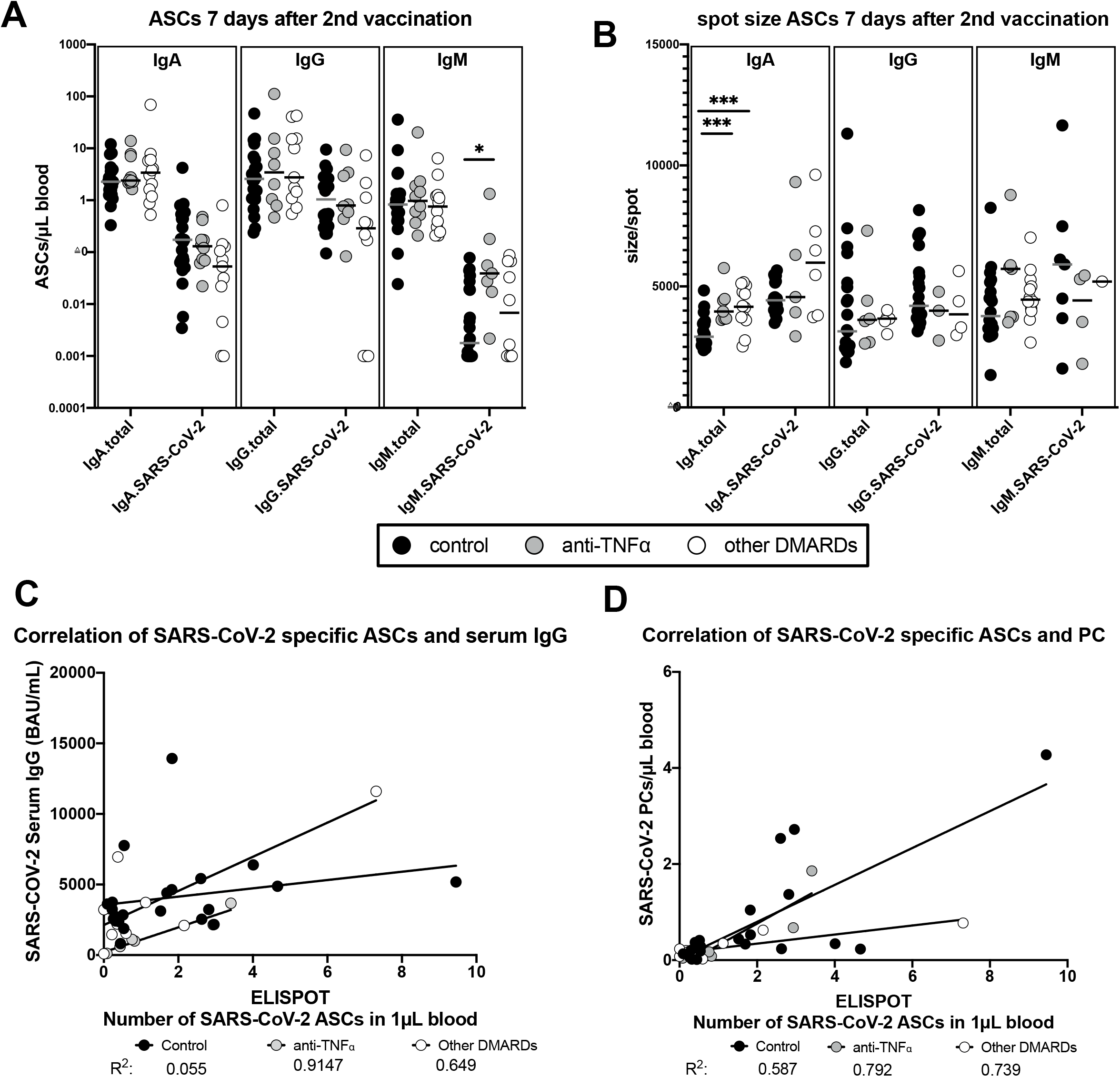
Antibody secreting cells (ASC) measured in 3-colour-Fluorospot 7 days after the second vaccination. See figure S3 for representative image. **A** Number of total and SARS-CoV-2 specific ASCs per µL blood. **B** Spot size distribution. Single points resemble the mean spot size per spot of one donor. Correlation (Pearson) of the number of SARS-Cov-2 specific ASCs **C** against SARS-CoV-2 specific plasma blasts in flow cytometry and **D** against SARS-CoV-2 serum IgG. Statistical differences: Kruskal-Wallis test with Dunn’s post hoc test, significant differences are indicated as *\*P<0*.*05, ***P<0*.*001*

### SARS-CoV-2 specific T cells show signs of delayed activation

SARS-CoV-2 S-specific CD4+ T cells could be detected at similar frequencies in all groups after second vaccination (figure 4A). We also observed no differences in cytokine production (TNFα, IFNγ, IL-2, IL-21 or IL-10) by the S-specific T cells between TNFα patients, oDMARDs and controls (figure 4B). In contrast, TNFα patients showed significantly lower expression of PD-1 and a trend towards increased levels of Ki-67, suggesting a delayed or still ongoing activation of these cells (figure 4C).

**Figure 4:**
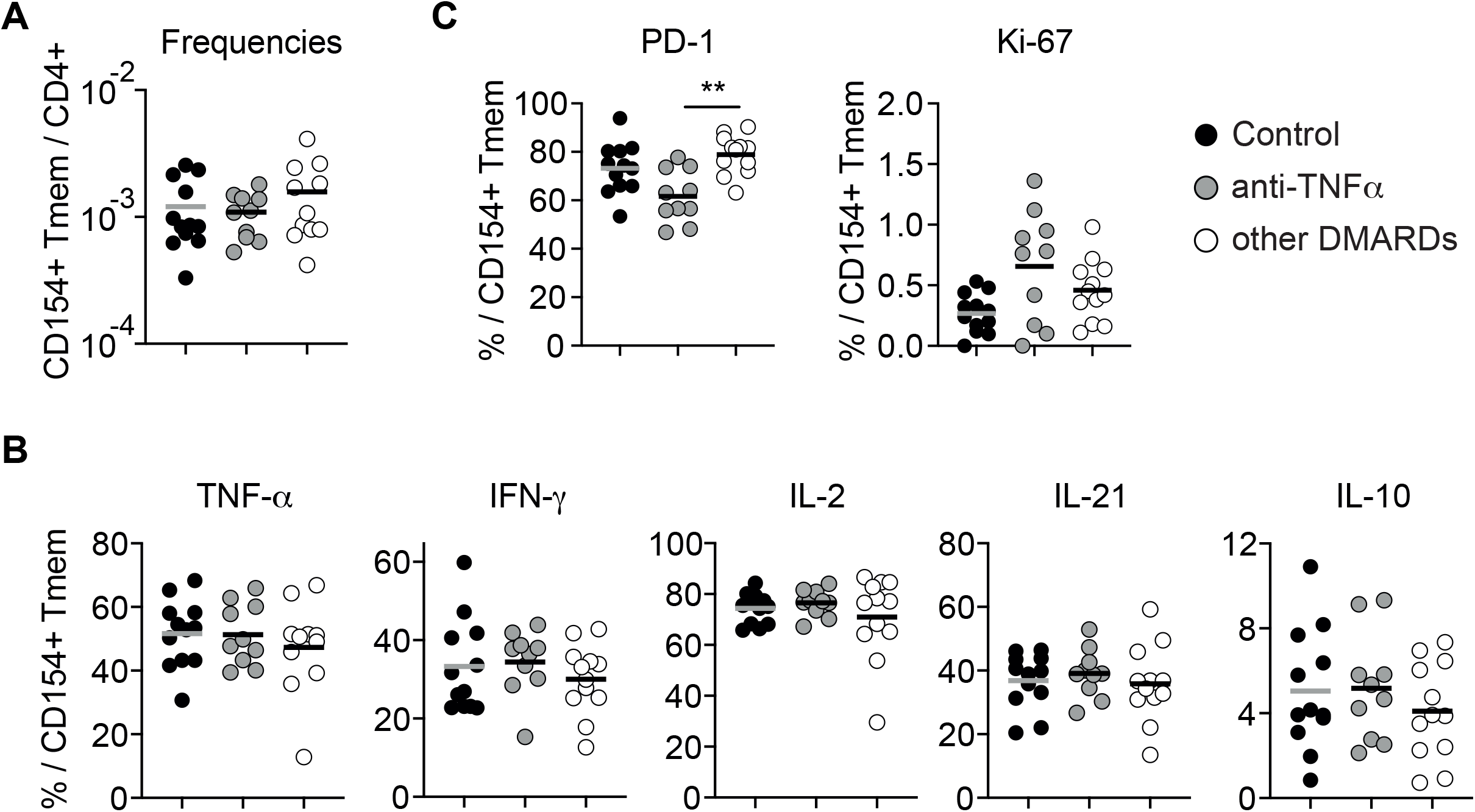
SARS-CoV-2 specific memory T cells in patients receiving TNFα blockers show a delayed activation status. Antigen-reactive T cell enrichment was performed two months after the second vaccination. **A** Frequencies of spike-reactive CD154+ memory cells within CD4+ T cells. **B** Percentage of cytokine production within spike-reactive CD154+ memory cells. **C** Percentage of PD-1 and Ki-67 positive cells within CD154+ memory cells. Each dot represents one donor, lines indicate mean values. Statistical differences: Kruskal-Wallis test with Dunn’s post hoc test, significant differences are indicated as *\*\*P<0*.*01*

## Discussion

To our knowledge, we present the first such comprehensive data on the longitudinal course of adaptive immunity in CID patients vaccinated against SARS-CoV-2 undergoing TNFα blockade.

These patients show an altered immune response after vaccination relative to oDMARD patients and healthy controls in the absence of breakthrough infections, suggesting that such individuals should be monitored more closely for loss of SARS-CoV-2 immunity.

Most strikingly, anti-TNFα patients exhibited a stronger decrease in IgG avidity and neutralisation capacity 6 months after vaccination and did not acquire NAs against BA.2 after a third vaccination. To our knowledge this is the first report concerning the long-term neutralisation efficacy against BA.2 VOC within this patient population after vaccination. While the neutralisation efficiency against the initial pre-VOC wt strain was marginally lower in anti-TNFα-treated patients 14 days after second vaccination compared to the other groups, this difference was more pronounced against the BA.2 strain where overall low neutralisation was detected. Six months after second vaccination, BA.2 NAs were not detected in any of the three study groups, suggesting that the S antigen of this VOC is relevantly different from that of wt and derived VOCs, consistent with recent studies.(10, 24) The loss of IgG avidity was unexpected as anti-TNFα patients displayed similarly high IgG avidity as the other groups at day 14 post second vaccination. Conversely, other studies have shown, that the avidity of anti-SARS-CoV-2 IgG increases during the subsequent months after vaccination.(13) This decline of avidity several months after the vaccination anti-TNFα patients has not been previously reported.

In addition, various changes in the plasmablast compartment of patients were observed in response to vaccination. Hence, patients using TNF inhibitors had higher numbers of plasma cells after the first vaccination (d0.2) relative to the two other groups. These differences increase after the second vaccination, suggesting a stronger immune reaction. The frequency of SARS-CoV-2 specific plasma cells within this population was decreased in the patient groups compared to controls. The absolute number of SARS-CoV-2 specific plasmablasts however was comparable to the other groups, suggesting that the immune reaction triggered under TNFα therapy is more unspecific.

Higher numbers of mature CD138^+^ plasmablasts were detected within the non-SARS-CoV2-specific plasma cells from anti-TNFα patients. These cells are usually found in the bone marrow.(25) The difference in maturity between S-specific and non-specific plasma cells may represent a reduced capacity to form long-lived specific plasma cells in anti-TNFα patients. However, assessment of these cells in the peripheral blood might not reflect their state in the bone marrow and our measurements after second vaccination may be too early for the detection of long-lived SARS-CoV-2-specific plasma cells. We also noted that a large proportion of these CD138^+^ plasma cells expressed HLA-DR (Figure S4). These phenotypically mature (CD138^+^) and antigen presenting plasmablasts (HLA-DR) were also described in patients with severe COVID19 (26) their function remains unclear.

Interestingly, although broadly homogeneous, there were two outliers in our cohort of anti-TNFα patients. One patient had consistently high antibody concentrations, high IgG avidity and NAs at 6 months. The other patient had consistently low values for all three parameters. These outliers show the importance of individual monitoring of patients at risk of reduced immunity after vaccination.

Our data demonstrate that TNFα inhibitors affect the adaptive immune response after SARS-CoV-2 vaccination. This is reflected in the development of IgG avidity, virus neutralising capacity, plasma cell and T cell populations. We recommend regular measurement of anti-SARS-CoV-2 IgG levels and especially NA titres in these patients, as this group may benefit from early booster vaccination. To our knowledge, current commercial anti-SARS-CoV-2 antibody tests are based on antigens still derived from the wild type. However, if available, assays adapted to the currently circulating variants should preferably be used. In addition, the development of variant-specific surrogate neutralisation tests would be desirable, as these, unlike live virus neutralisation assays, can also be used in routine laboratories. CID patients on TNFα inhibitor therapy who have no, or low detectable antibody levels should be particularly protected from COVID-19. Following SARS-CoV-2 infection, these patients might require close monitoring and early administration of monoclonal antibodies that also cover currently circulating VOC. In addition, we recommend the use of a vaccine adapted to the current VOC as soon as it becomes available.

The mechanisms leading to decreased antibody response during immunosuppressive treatment need to be further explored to improve vaccine regimens for these high-risk patients.

## Data Availability

All data produced in the present study are available upon reasonable request to the authors

## Contributors

Study design: UMG, FT, MP, SS, AK, HF, BFH. Sample collection: DKB,ACL, JHS, MS, FT, RZ, SG. Experiments and data analysis: UMG, RR, FN, MC, LV, HMR, PB, FB, MH, MV, AK, HF and BFH. Tables and figures: UMG, RR and BFH. Data interpretation: UMG, RR, PB, AK, HF and BFH. Writing of the manuscript: UMG, PH, PJM, AK and BFH. Critical proof reading of the manuscript: all authors.

## Acknowledgement

We thank Meike Zahnen (UKSH, Rheumatology), Ina Martens (UKSH, Rheumatology), Sina Müller (Labor Dr. Krause und Kollegen) and Carina Bäumler (Labor Dr. Krause und Kollegen) for excellent technical and organisational assistance.

We acknowledge the support of the European Union for the ATAC (‘‘antibody therapy against corona’’. Horizon2020 number 101003650) consortium.

The authors thank Mikrogen GmbH, Neuried, Germany, for providing free or discounted kits for IgG avidity detection.

