## Supplements for "Multiple effects of TNFα inhibitors on the development of the adaptive immune response after SARS-CoV-2 vaccination"

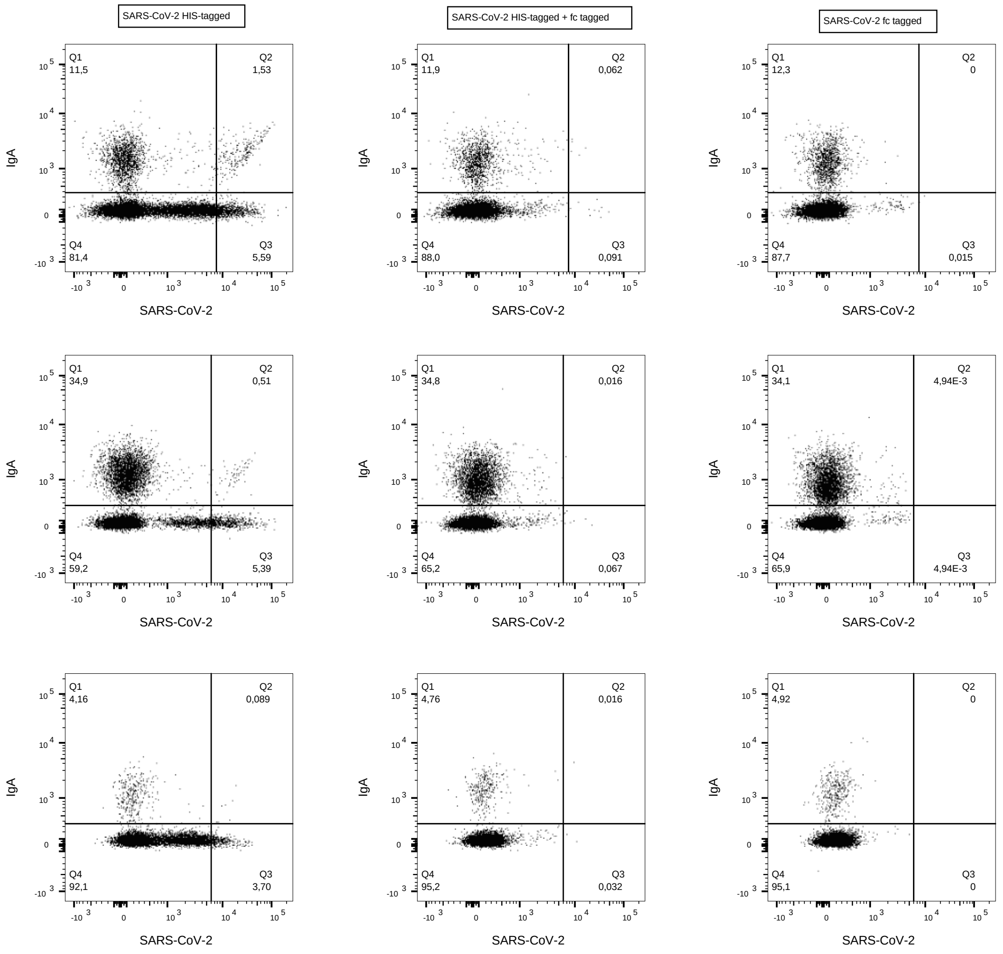


*Figure S1:* 3 Gating strategy with blocking control for three representative samples. PBMCs were isolated, stained and plasma cells were gated as described before. Samples from the same donor were incubated with either his-tagged S1-Protein (left), fc-tagged S1-protein (right) or both (middle) before staining. Data of three representative individuals is shown. This gating, used for each patient in the study, was adjusted according to figure S2 to exclude false positive cells. Therefore, the gating seems to cut through the positive population.

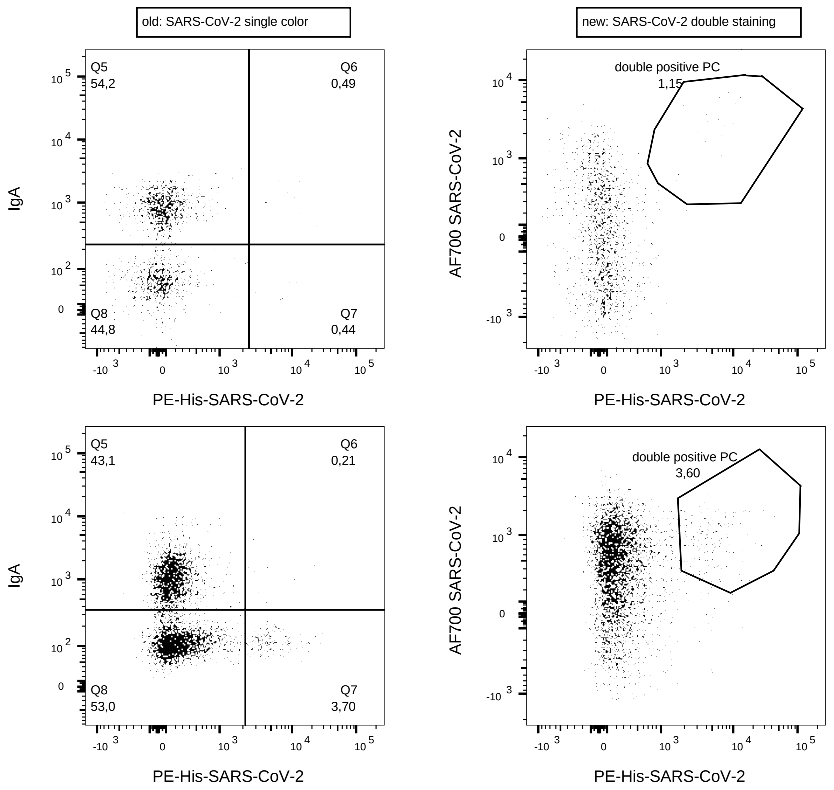


*Figure S2:* Comparison of the PE-HIS-SARS-CoV-2 single staining used in the study with a double staining. Before the experiment, a fc-tagged version of the S1 protein was labelled with AlexaFluor700 (Abcam, Cambridge, UK) which was incubated together with the His-tagged S1 protein before staining with the B cell antibody cocktail which included the PE-His antibody. Here we show two representative individuals. The gating used for all patients in the study was adapted according to the figures on the left-hand side and figure S2.

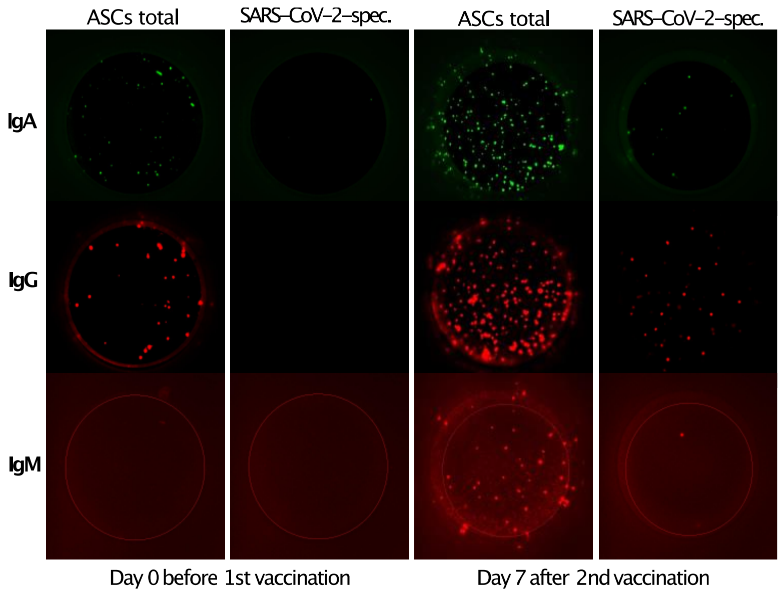


*Figure S3: Fluorospot data 7 days after the second vaccination. Representative image of $3*10^5$ PBMCs per well.*

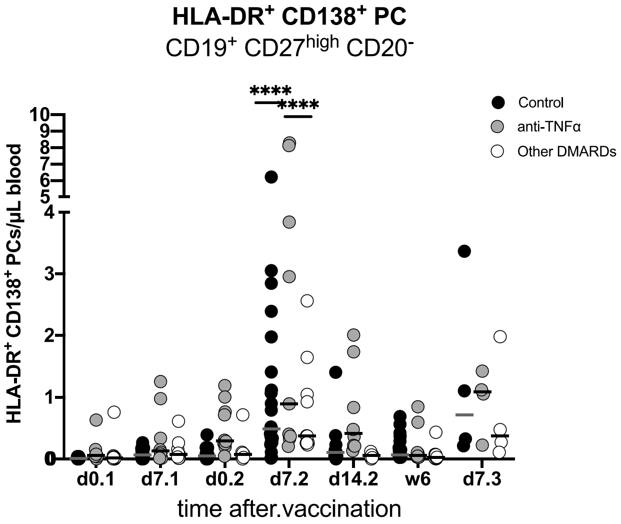


*Figure S4: mature activated HLA-DR/CD138 double positive plasma cells. Cell counts per microlitre of blood are shown as single values with median. Statistical differences: Kruskal-Wallis test with Dunn's posthoc test, significant differences are indicated as ****P<0.0001*
